# Modelling the impact of vaccination on SARS-CoV-2 transmission in England

**DOI:** 10.1101/2024.09.06.24313210

**Authors:** Nieves Derqui, Swapnil Mishra, Wes R Hinsley, Samir Bhatt, Daniel J Laydon

## Abstract

While the efficacy of SARS-CoV-2 vaccination against severe disease and mortality is well-established, its impact on population-level transmission remains a critical yet poorly quantified frontier in public health. Previous evidence has largely relied on small-scale cohort or household studies. However, these settings often lack the scale to capture the broader ecological impact of vaccination on epidemic growth, and therefore its implications for epidemic control.

Here, we quantify the effectiveness of SARS-CoV-2 vaccination against transmission in England by fitting a Bayesian hierarchical model to estimates of the time-varying effective reproduction number (*R_t_*) during 2021. Vaccine effectiveness for one, two and three doses was defined as the proportional reduction in *R_t_*. The model integrates high-resolution data on vaccination uptake by age group, the shifting landscape of circulating variants, and regional transmission trends across 221 Lower Tier Local Authorities.

We find that a first vaccine dose provided moderate-to-large effectiveness against transmission (45.4% (95% CrI: 38.5–51.9%)), whereas the second dose offered negligible additional transmission-blocking effects. A third dose significantly restored effectiveness (66.3% (40.3–90.1%)), albeit with higher uncertainty. Surprisingly, although a link between socio-economic deprivation and transmission is highly plausible, we find that deprivation was not a significant driver of transmission heterogeneity during the vaccination rollout. The model accurately reproduces observed spatial and temporal variation in *R_t_* across England.

To our knowledge, these findings provide the first population-level evidence that vaccination substantially reduces the SARS-CoV-2 reproduction number, supporting the UK’s “first-doses-first” prioritization as effective tools for epidemic control. More broadly, our modelling framework offers an approach for assessing transmission-blocking effects of vaccines against respiratory pathogens using routinely collected surveillance data, and has implications beyond SARS-CoV-2 for emerging threats such as avian influenza (H5N1).

## Introduction

Vaccination is a key intervention for the control of respiratory viruses, yet vaccination effectiveness against transmission remains poorly quantified. SARS-CoV-2 vaccines were proven safe and efficacious in clinical trials (1-3), and their effectiveness was subsequently demonstrated against infection, symptomatic disease, hospitalisation and death (4, 5). SARS-CoV-2 vaccines also proved efficacious against the Alpha and Delta variants (6), and to a lesser extent against Omicron. Vaccination is estimated to have averted 14.4 million SARS-CoV-2 deaths globally (7).

However, the impact of vaccination on SARS-CoV-2 transmission is less understood (8, 9), with most evidence coming from household cohort studies that analyse onward transmission from index cases (10, 11). While these studies have unique advantages, such as capturing incident infections, they are often limited by small sample sizes and to specific settings. To our knowledge, no previous study has quantified vaccine effectiveness against SARS-CoV-2 transmission at the level of a population. Knowing this effectiveness is important for public health planning, as it informs not only a vaccine’s direct effects in reducing individual risk of hospitalization and death, but also its indirect effects on a population in preventing onward transmission.

In the UK, SARS-CoV-2 transmission varied markedly between regions and over time (12), due to: regional differences in viral variant spread (13-16); the stringency of, and adherence to, non-pharmaceutical interventions (17); and vaccine uptake (18). This regional variation presents a difficulty in accurately modelling SARS-CoV-2 transmission dynamics and the effects of vaccination upon them.

However, this same regional variation presents an opportunity, because differences between regions in their vaccination uptake can be compared to differences between regions in their transmission, in effect providing a spatial natural experiment. Here, we develop a Bayesian hierarchical model and apply it to high-resolution data from England available at Lower Tier Local Authorities (LTLA) level. The model leverages several such data sources: i) routinely collected data on vaccination uptake by age, LTLA and dose from the UK Health Security Agency (UKHSA); ii) the proportion of circulating variants from the COVID-19 Genomics UK (COG-UK) Consortium; and iii) matched regional estimates of the time-varying effective reproduction number (*R_t_*) for 221 LTLAs.

Our framework is not pathogen-specific, and can serve as a template for evaluating the impact of vaccination against other respiratory threats, such as H5N1 or seasonal influenza, where spatial and temporal heterogeneities exist. By leveraging these spatial heterogeneities, our approach demonstrates how routinely collected surveillance data can be utilized to infer the population-level effectiveness of vaccination.

## Methods

### Data sources and definitions

Vaccination data was available from UK Health Security Agency (UKHSA) (19) from 8^th^ December 2020 until 14^th^ November 2021 (Figure 1A). The number of vaccinated individuals was available, aggregated for all age groups and per five-year age bands, for every Lower Tier Local Authority (LTLA) at each timepoint.

**Figure 1:**
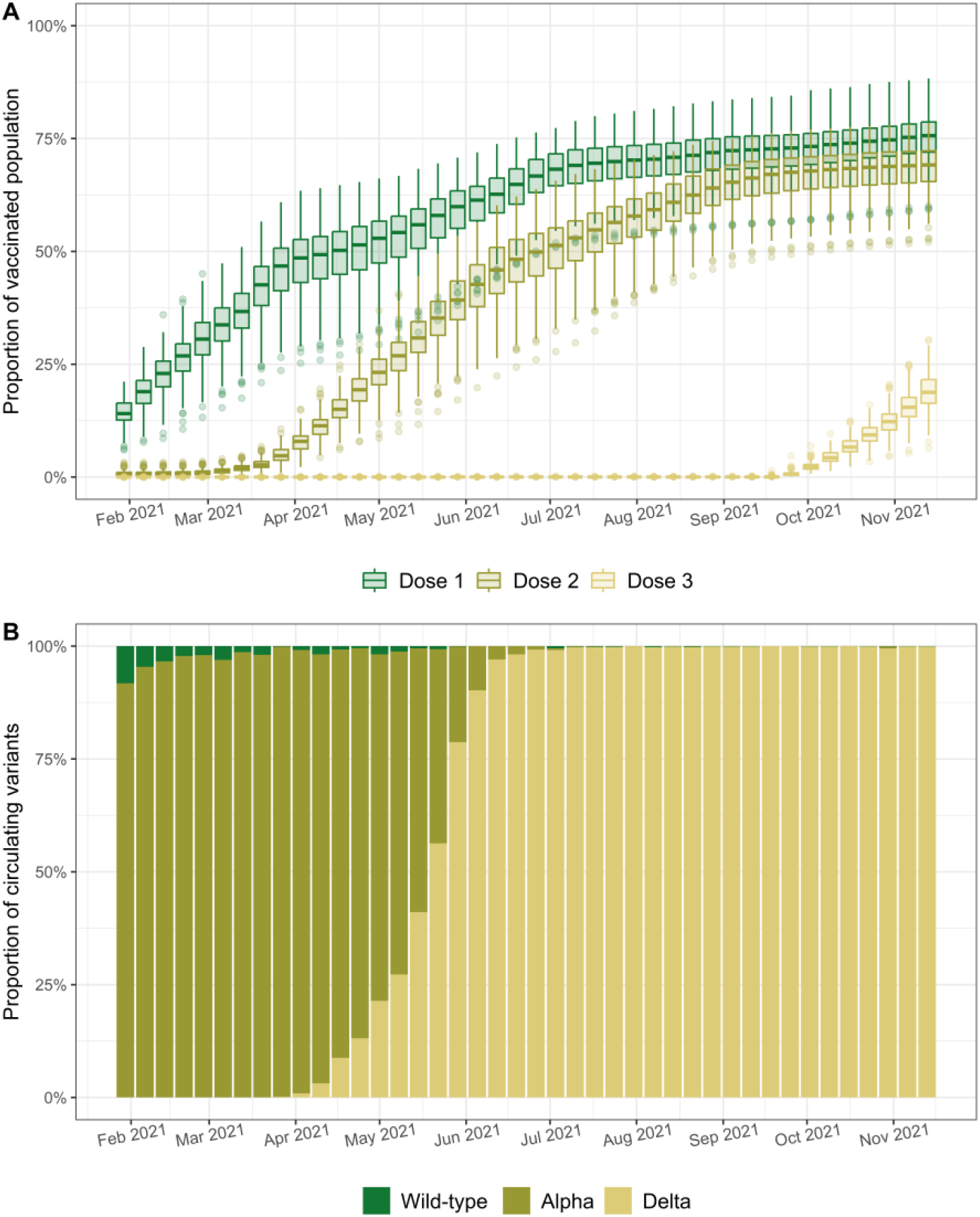
A) Time-series of boxplots showing vaccine uptake by dose across England in 2021. B) Time-series of proportion of SARS-CoV-2 variants circulating across England in 2021. Boxplots represent the distribution (across 221 Lower Tier Local Authorities (LTLAs)) of the proportion of those vaccinated with one, two and three doses at each timepoint (with 1st quartile, median and 3rd quartile shown). Whisker limits correspond to maximum and minimum values, whenever these remained within 1.5xIQR range; otherwise, points outside this range are individually plotted as outliers. Abbreviations: IQR, Inter-Quartile Range.

Data on the proportions of “wild-type”, Alpha and Delta infections detected in each LTLA over time was calculated using the UKHSA’s variants and mutations (VAM) line list, as well as public genomic survey data provided by the COVID-19 Genomics UK (COG-UK) Consortium. These data were used to estimate each LTLA’s proportion of circulating “wild-type”, Alpha and Delta variants. Importantly, data was only available for these three variants (i.e. excluding Omicron), and thus the sum of the proportions of wild-type, Alpha and Delta variants for every LTLA and timepoint was normalised to one. Data on circulating variants was available from 2^nd^ February 2020 until 5^th^ July 2022 (Figure 1B).

We used estimates of the real-time effective reproduction number from a previously established method (20), which considered national and regional data on daily SARS-CoV-2 cases and deaths, together with sero-surveillance data. For brevity, we will henceforth refer to these estimates as the “observed *R_t_*”, even though this is a slight misnomer as *R_t_* cannot be directly observed. These observed *R_t_* estimates were available from 30^th^ January 2021 until 2^nd^ January 2022 for each timepoint and LTLA (Figure 2).

**Figure 2:**
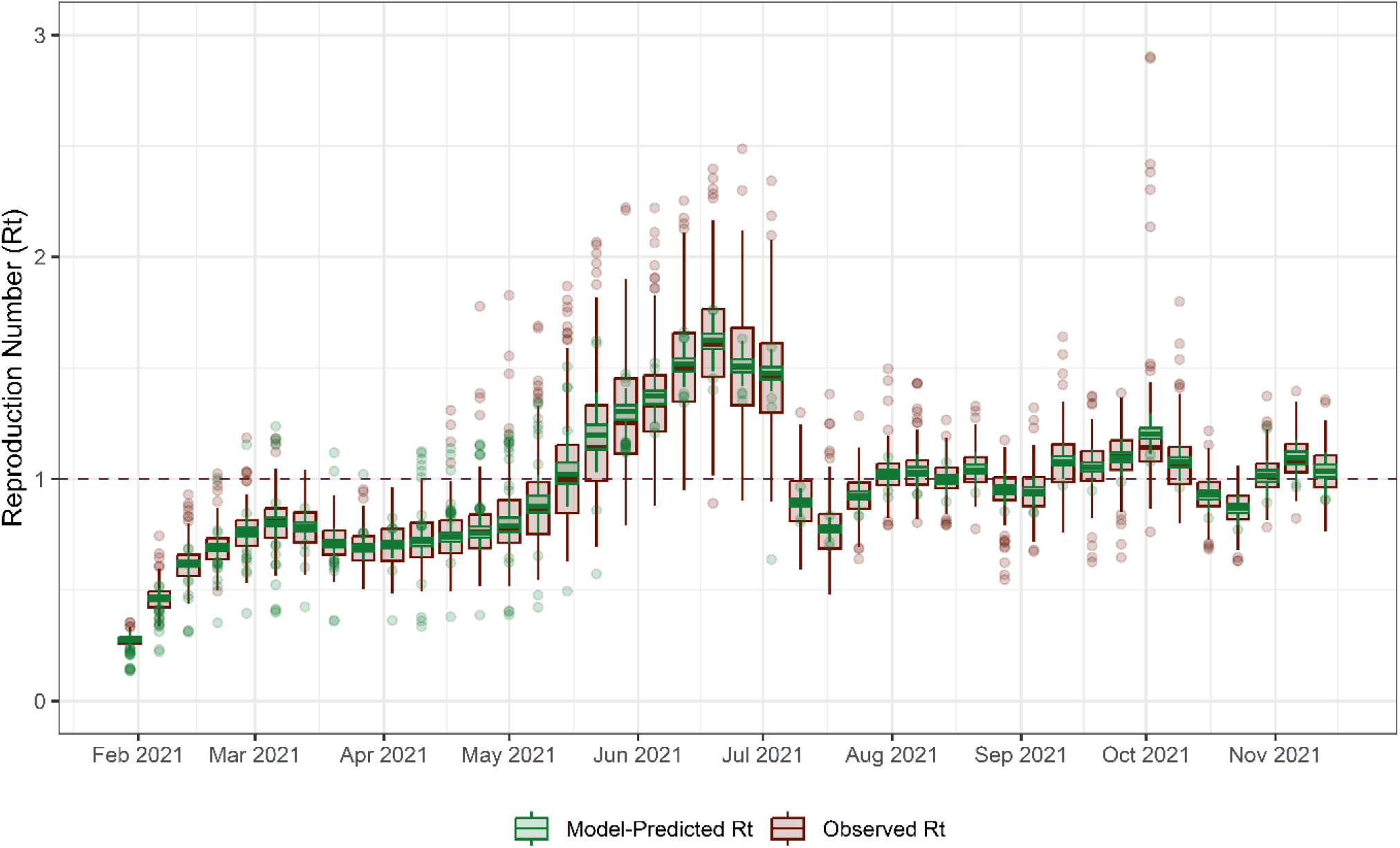
Time-series of boxplots showing observed *R_t_* and predicted *R_t_*. Boxplots represent distribution (across 221 Lower Tier Local Authorities (LTLAs)) of *R_t_* estimates at each timepoint (1st quartile, median and 3rd quartile shown). Whisker limits correspond to maximum and minimum values, whenever these remained within 1.5xIQR range; otherwise, points outside this range are individually plotted as outliers. Abbreviations: IQR, Inter-Quartile Range.

Only timepoints with complete data (i.e. where vaccination, circulating variants and observed *R_t_* data was available) were considered, and so our study period is 30^th^ January 2021 until 14^th^ November 2021. This precluded analysis of Omicron as it was only detected in the UK in late 2021 (21). The final dataset included 9,282 total observations from 221 LTLAs over 42 weeks.

### Model description

Let *λ*(*t*) be the baseline transmission trend (treated discretely in weeks), which is scaled by a factor *g*_*m*_ in each region *m*. So here, *λ*(*t*) and *g*_*m*_*λ*(*t*) respectively describe the trend of the SARS-CoV-2 reproduction number at national level, and at regional (LTLA) level, at time *t*, in the absence of vaccination, and without consideration of viral variants.

For variant *v* = 1, …, *N*_*v*_, let *p*_*vmt*_ be the proportion of variant *v* in LTLA *m* at time *t*. We define *M*_*v*_as the relative transmission advantage of variant *v*. Thus, the LTLA-specific trend at time *t* in LTLA *m* due to variant *v* (without vaccination) is given by *p*_*vmt*_*M*_*v*_*g*_*m*_*λ*(*t*). Considering only wild-type, Alpha and Delta variants (see above), we assume that 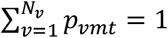 for all times *t* and all regions *m*. Note that a variant’s relative transmission advantage is relative to wild-type variant, and thus *M*_1_ = 1 by definition.

The reduction in transmission from dose *d* = 1, …, *D* is given by (1 − *ρ*_*mdt*_*VE*_*d*_), where *ρ*_*mdt*_ is the proportion of vaccinated individuals in each LTLA *m* at a timepoint *t* with who have had *d* doses, and where *VE*_*d*_ is the vaccination effectiveness against transmission in a population 100% vaccinated with dose *d*. A maximum of *D* = 3 doses was considered.

Therefore, the mean reproduction number *R*_*m*_(*t*) in LTLA *m* at time t is given by

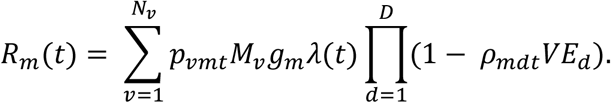

Finally, we allow the observed reproduction number 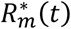 to be normally distributed around *R*_*m*_(*t*) with standard deviation *σ*, i.e.

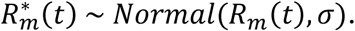

A glossary of parameters is given in Table 1, together with prior distributions for fitted parameters.

**Table 1:**
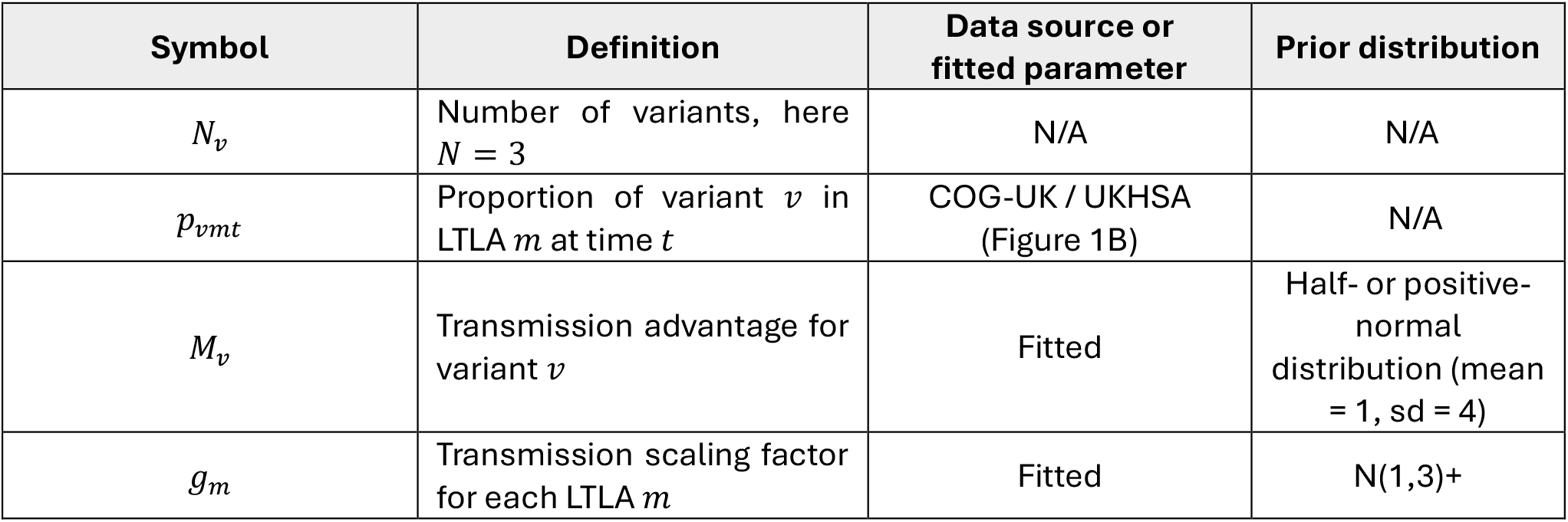
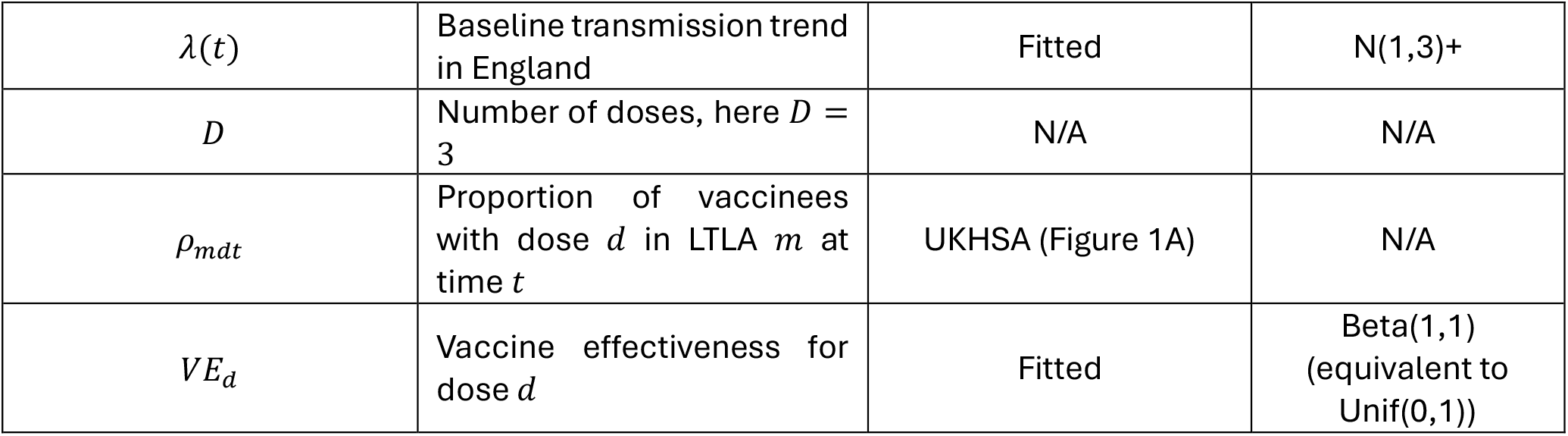
Model terms glossary. Data source is indicated for model inputs, and for terms fitted in the model, prior distribution is indicated.

We report the model-predicted *R_t_* for each LTLA and timepoint. Additionally, as a counterfactual, we estimated *R_t_* in the absence of vaccination 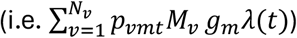. In all model-predicted estimates, we calculated the mean estimate across parameter iterations and the 2.5% and 97.5% credible intervals (CrI).

### Sensitivity analysis and model selection

We considered several alternatives to the main model presented above: a model in which vaccination effectiveness varied by variant as well as by dose, termed “Variant-specific-VE”; a model that used the age-group-proportion of vaccinated individuals but whose VE did not change per variant or age group, termed “Age-model”; and a model alternative in which vaccination effectiveness varied by dose, variant and age group, termed “Age-variant-specific-VE”. Further information on these models is detailed in the supplementary materials. Model selection was conducted using Leave-One-Out Cross-Validation (LOO-CV) (22).

### Spatial Heterogeneity and Deprivation Analysis

To investigate whether regional transmission was associated with socio-economic factors, we extended the baseline model to include the 2019 Index of Multiple Deprivation (IMD) (23). We utilised the “Average Score” across all domains of deprivation, aggregated at the LTLA level. For the 221 LTLAs included in the study, the IMD scores were z-score standardised (mean = 0, standard deviation = 1) to allow the estimated effect size to be interpreted as the proportional change in *R*_*m*_(*t*), per one standard deviation increase in deprivation.

We formulated the deprivation effect as a multiplicative term exp (*ζ* · *IMD*_*m*_) applied to the regional scaling factor *g*_*m*_. This approach ensures that the impact of deprivation is proportional to the local *R*_*m*_(*t*) and maintains the requirement for non-negative transmission rates. The total spatial multiplier for LTLA *m* was therefore *g*_*m*_exp (*ζ* · *IMD*_*m*_), giving

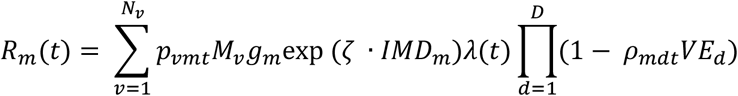

where *ζ* ∼ *N*(0,0.5) is the estimated coefficient for the deprivation effect. Note that our main model is nested within this IMD model, and thus a *ζ* value of zero would indicate that deprivation has no impact on *R*_*m*_(*t*), while positive or negative values indicate increased or decreased transmission potential associated with higher deprivation, respectively.

### Software

Analyses were conducted using R version 4.2.3 (24). The model was implemented and fitted using Hamiltonian Monte Carlo in Stan (25), through the R package ‘*rstan’* (26), using 10,000 iterations with a warm-up of 2,500 iterations across 10 chains. Leave-One-Out Cross-Validation was performed with the ‘loo’ package (27). All plots were generated using ‘ggplot2’ (28).

### Data availability

All model code is available at https://github.com/NDerqui/COVID_LTLA_Rt_Paper.git.

## Results

### SARS-CoV-2 vaccination uptake and circulating variants

The proportions of people vaccinated with one, two or three doses across all LTLAs during the study period is shown in Figure 1A. Vaccination with dose one commenced across England in January 2021, followed by dose two in April. By mid-April, the median proportion of individuals vaccinated with at least one dose surpassed 50% across LTLAs, while dose two reached the 50% median threshold in July. Variability in uptake between regions remained high throughout the study period, particularly during the plateau observed in autumn 2021 where dose one and dose two coverage overlapped significantly. Rollout of the third (booster) dose began in October as combined coverage for the first two doses approached 75%.

The proportions of circulating variants are depicted in Figure 1B. The study period was characterised by the sequential dominance of the Alpha and Delta variants (15, 29). While the wild-type variant was still circulating at low numbers in late January 2021, by late March, Alpha was dominant, to itself be rapidly displaced in April by the Delta variant. Delta achieved total dominance by July 2021 and remained the primary variant until the introduction of Omicron late in the year, with a few small and sporadic Alpha variant appearances.

### *R_t_* evolution and modelling results

The model effectively reproduced the observed regional and temporal variation in *R_t_* across all 221 LTLAs (Figure 2). Observed *R_t_* estimates remained below 1 from January until April, coinciding with national lockdown measures. During this period, *R_t_* was around 0.6, with little variability between LTLAs. A gradual increase in *R_t_* followed, with median values rising from 0.7 in April to 1.5 in July as restrictions eased and the Delta variant expanded. At this point, variability across LTLAs was more pronounced, with maximum *R_t_* values reaching 2.5. Following a sharp decline in mid-July, *R_t_* stabilized near 1 for the remainder of the study period, with less regional variability. Our model fitted the observed *R_t_* values well (Figure S1), and effectively reproduced the regional and temporal variation in the reproduction number across all 221 LTLAs over time (Figure 2).

Modelling fits for nine example LTLAs are depicted in Figure 3, and fits for the remaining LTLAs are shown in Figures S2-S12. The model reproduces the individual LTLA level trends in the *R_t_* well across the 221 LTLAs, although there are some interesting differences across a minority of LTLAs. For example, the peak in July is earlier and much higher among the observed *R_t_* than the model-predicted in Liverpool (Figure 3), while in Westminster the model predicted one single peak when there were in fact three smaller peaks (Figure S12). In general, the model’s *R_t_* trends are more consistent across LTLAs than are the observed *R_t_* trends.

**Figure 3:**
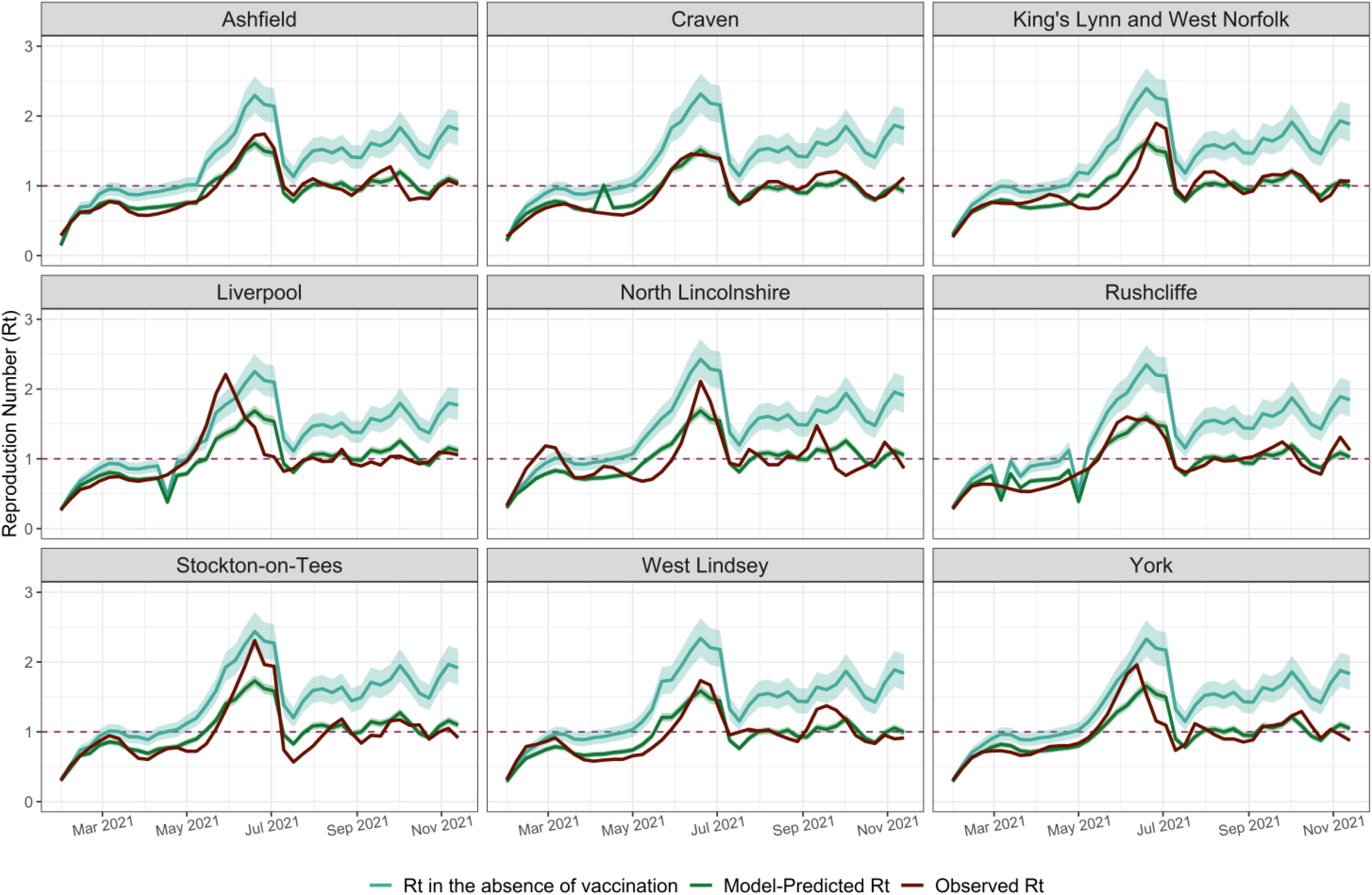
Model fits in nine example LTLA: *R_t_* in the absence of vaccination (counterfactual), model-predicted and observed *R_t_* in 2021. Each LTLA *R_t_* in the absence of vaccination (counterfactual), model-predicted *R_t_* and observed *R_t_* are plotted as a continuous line. The 2.5% and 97.5% percentiles for each LTLA *R_t_* in the absence of vaccination and model-predicted *R_t_* are plotted as a ribbon line. Abbreviations: LTLA, Lower Tier Local Authority; *R_t_*, Reproduction Number.

At the beginning of 2021, the counterfactual *R_t_* values that represent transmission in the absence of vaccination, are highly similar to the observed *R_t_* (Figure 3), due simply to the low proportions of vaccine uptake at that time. By July, when most people had received at least one dose, the discrepancy between the counterfactual and observed *R_t_* became much wider (roughly, counterfactual and observed *R_t_* were 2 and 1.5, respectively). Consistent with the plateauing of dose 1 uptake at this point, this discrepancy between the counterfactual and observed *R_t_* stopped widening then, and by the end of the year, individual LTLA counterfactual *R_t_* trends had values around 1.5 while observed and model-predicted *R_t_* stayed close to 1.

We also estimated the extent to which vaccination reduced *R_t_* overall across all LTLAs. Average across all timepoints and LTLAs of our predicted *R_t_* estimates was 0.9633, and average across all timepoints and LTLAs of our counterfactual *R_t_* was 1.3665, meaning we estimated an overall 29.51 % reduction of *R*_t_ by vaccination. However, average predicted *R_t_* and counterfactual *R_t_* across all LTLAs up until the end of April were 0.67 and 0.81, respectively, and average predicted *R_t_* and counterfactual *R_t_* across all LTLAs up until mid-July were 0.93 and 1.23, respectively. Thus, the overall effect of vaccination until the end of April was 17.3% (when median proportion of dose one vaccinees surpassed 50%) and 24.2% by mid-July (when median proportion of dose two vaccinees surpassed 50% but median proportion of dose one vaccinees was close to 75%), reflecting little effect by dose two vaccination.

Our counterfactual is imperfect, in that other measures would likely have been implemented (e.g. greater social distancing, whether mandatory or voluntary) had there genuinely been no vaccine, but this is nevertheless informs the population-level transmission impact of vaccination.

### Effect of vaccination on transmission

Dose one demonstrated moderate effectiveness against transmission at 45.4% (95% Credible Interval: 38.5% - 51.9%) (Figure 4A). In contrast, dose two had a negligible incremental effect on transmission. The largest vaccine effectiveness against transmission was estimated for dose three, 66.3% (40.3% - 90.1%), but this estimate also had the widest credible intervals, due to the limited duration of the booster rollout within the study window.

**Figure 4:**
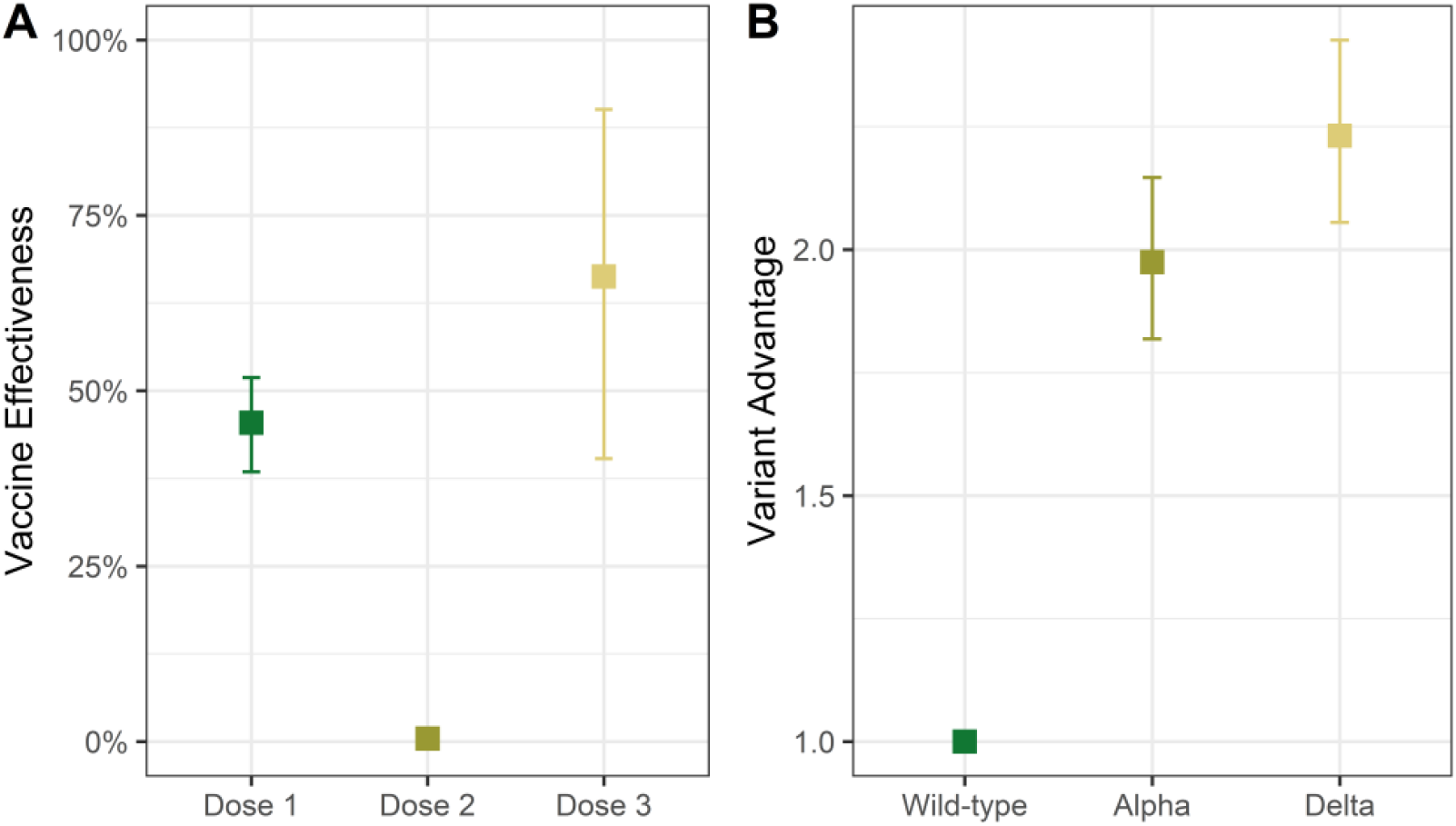
A) Effectiveness of vaccination against transmission by dose. B) Variant advantage. Model predictions for vaccine effectiveness (A) and variant’s advantage (B) are shown as point estimates depicting the average across iterations, with error bars showing 2.5% percentile 97.5% percentile.

The model further estimated the transmission advantage of viral variants relative to the wild-type. The Alpha variant was found to be 2.0 (95% CrI: 1.8–2.2) times more transmissible than pre-Alpha variants, while the Delta variant exhibited a 2.2 (95% CrI: 2.1–2.4) transmission advantage (Figure 4B).

### Model selection

We considered several alternatives to our main model, that allowed vaccine effectiveness to vary by age, or by SARS-CoV-2 variant. Our estimates of effectiveness are largely robust to model variant, with similarities and differences discussed below.

Vaccine effectiveness estimates from the “Age-model” were very similar to those of the main model (51.6% (40.7% - 60.8%), 3.8% (0.1% - 12.9%), and 40.9% (2.3% - 92.0%) for doses one, two and three) (Figure S13). Although the “Variant-specific-VE” model showed considerably higher dose one effectiveness estimates for wild-type and Alpha variants (97.6% (91.4% - 99.9%) and 82.7% (76.5% - 88.5%), respectively), it is noteworthy that as dose one coverage increased, the proportion of these variants decreased. Thus, these vaccine effectiveness estimates may be an artefact of the model. This model variant also estimated very high dose two effectiveness for wild-type variants (75.4% (33.6% - 98.9%) but with high uncertainty. The final model, “Age-variant-specific-VE”, estimates had uncertainty that was too great to draw meaningful conclusions (Figure S13).

Models without age-group proportion of vaccinees showed a better fit than those with age-group proportion of vaccinees (Table S1), as they had higher ELPD. The “Variant-specific-VE” model yielded a higher Expected Log Predictive Density (ELPD), suggesting a superior fit to the observed data. However, we favour the Main Model for several structural and epidemiological reasons. While the variant-specific version adds only six additional effectiveness parameters, these are “global” parameters that influence the transmission of all 221 LTLAs simultaneously. In a Bayesian hierarchical framework, such global parameters exert significantly more leverage than local LTLA-specific scaling factors. Because the disappearance of the Alpha and the wild-type variants coincided precisely with the peak of dose one administration, these global efficacy parameters are susceptible to temporal correlation artefacts. Specifically, the model cannot cleanly distinguish between a variant’s disappearance due to high vaccine effectiveness or its displacement by a more fit emerging variant. Consequently, the Main Model provides a more robust and conservative estimate of vaccine effectiveness, less susceptible to these identifiability issues.

Nevertheless, the estimates produced by the “Variant-specific-VE” model largely validate the conclusions of the Main Model, as the estimates of efficacy by dose are consistent (Figure S13). For dose one, the Main Model’s ∼45% pooled effectiveness is consistent with the variant-specific weighted average (high for wild-type and Alpha, lower for Delta). For dose two, both models confirm near-zero transmission effectiveness against Alpha and Delta. High point estimates for wild-type are epidemiologically meaningless, as dose two occurred with virtually no circulating wild-type and therefore estimates exhibit extremely wide credible intervals. For dose three, both models show moderate-to-high effectiveness against Delta, while producing non-informative, wide intervals for the then-extinct wild-type and Alpha variants.

We conclude that the Main Model’s pooled approach offers a parsimonious and epidemiologically grounded representation of vaccine impact. The consistency of results for the primary circulating variants across both models further provides confidence in the robustness of the Main Model’s findings.

### Spatial Heterogeneity and Deprivation Analysis

To evaluate the extent to which socio-economic factors drove regional differences in transmission, we assessed the relationship between the Index of Multiple Deprivation (IMD) and the reproduction number *R*_*m*_(*t*), across 221 LTLAs.

Surprisingly, this extension of our main model revealed that deprivation was not a significant predictor of transmission potential during the study period. The estimated deprivation effect size *ζ*= -0.012 (-0.112 – 0.079) was negligible and tightly centred around zero. Therefore, this extended model essentially reduces to our (nested) main model.

Crucially, the inclusion of IMD did not alter the primary findings regarding vaccine effectiveness. The estimated effectiveness for one, two, and three doses remained extremely consistent between the main and deprivation models (e.g. dose one: 45.4% vs 45.4%; dose three: 66.3% vs 66.4%). This stability demonstrates that the observed spatial heterogeneity in *R*_*m*_(*t*), is likely driven by factors other than socio-economic deprivation, such as stochastic variant introduction and local contact networks, and that our primary estimates of vaccine impact are not confounded by LTLA-level deprivation scores.

## Discussion

Despite the well-established benefits of vaccination against SARS-CoV-2 infection, severity and death, our understanding of the impact of vaccination on transmission at population level remains limited. In this study, we developed a Bayesian hierarchical model to estimate the effectiveness of vaccination on SARS-CoV-2 transmission in England over 2021, quantifying the reduction on the effective reproduction number (*R_t_*) as population-level outcome. We found that a first vaccine dose provided moderate-to-large effectiveness against transmission (45.4%, 38.5% - 51.9%), while the third dose restored protection (66.3%, 40.3% - 90.1%). Notably, our estimates suggest that the second dose provided negligible additional transmission-blocking benefits beyond those already achieved by the first.

The model accurately reproduced the temporal evolution of the reproduction number across 221 LTLAs, capturing the rise in transmission potential during spring 2021 as social restrictions were eased (30) and the more transmissible Delta variant emerged (31, 32). This lends support to our hypothesis, namely that vaccination was a major determinant of transmission intensity.

However, the model initially predicted less spatial variability across LTLAs than was observed, prompting us to include the Index of Multiple Deprivation (IMD) as a covariate. However, perhaps counteractively, we found that this did not affect our estimates of effectiveness as the effect size of IMD was essentially zero. This suggests that during the 2021 rollout, the transmission landscape was dictated more by viral evolution and vaccine-induced immunity than by socio-economic factors. This finding highlights the robustness of our primary results, as the estimated impact of vaccination remained stable even when accounting for a highly plausible socio-demographic confounder.

Our estimate for dose one effectiveness (45.4%) is broadly consistent with cohort studies, such as those among healthcare workers in Cambridge (75% reduction in infection) (33) and residents in long-term care facilities (56% reduction) (34). While our estimates are lower than some household-based studies, which have reported effectiveness as high as 61% and 66% after vaccination with one dose of ChAdOx1 and BNT162b2 respectively (11), this discrepancy likely reflects our use of *R_t_*, a community-wide measure, versus outcomes limited to specific high-contact settings. Clifford et al. estimated a dose two effectiveness of 36% (-1%, 66%) for BNT162b2 and 49% (18%, 73%) for ChAdOx1, which are more similar to our dose one estimates (10). However, even after accounting for the concurrent rise of the Delta variant and its increased transmissibility, our estimates still suggest that dose two had a negligible additional benefit.

This finding supports the “first-doses-first” policy adopted in the UK (35, 36). By prioritizing rapid first-dose coverage, the programme may have achieved a saturation of transmission-blocking effects across the population before the second doses were administered. It is important to note that our model accounts for dose effects cumulatively; the null result for dose two indicates no *marginal* gain in transmission reduction, which is consistent with some studies that described only a mild difference between vaccination with one and two doses (5).

The moderate-to-high effectiveness estimated for the third dose (66.3%) highlights the importance of boosters in maintaining epidemic control. Although this estimate carries substantial uncertainty due to the limited overlap between the booster rollout and the rise of the Omicron variant, it aligns with a restoration of immunity following the well-documented waning of early doses.

Our study has limitations. We assumed constant vaccine effectiveness over the study period, thus not explicitly modelling the waning of protection over time (37). Furthermore, while we attempted to include age-specific vaccine effectiveness, the resulting credible intervals were non-informative, likely indicating that the model was underpowered to resolve these parameters at the LTLA level.

To our knowledge, this study provides the first population-level evidence that vaccination substantially reduces the SARS-CoV-2 reproduction number. Our framework offers a scalable, robust approach for assessing the transmission-blocking effects of vaccines using routinely collected surveillance data. This methodology has implications beyond the current pandemic, offering a template for evaluating interventions against future respiratory threats, such as avian influenza (H5N1), where quantifying impact on community-level transmission will be paramount for epidemic control.

## Data Availability

All model code is available at https://github.com/NDerqui/Vaccine_Model.

## Other information

## Acknowledgments

ND, WRH, SB and DJL acknowledge funding from the MRC Centre for Global Infectious Disease Analysis (reference MR/X020258/1), funded by the UK Medical Research Council (MRC). This UK funded award is carried out in the frame of the Global Health EDCTP3 Joint Undertaking. SM acknowledges support from the National Research Foundation via The NRF Fellowship Class of 2023 Award (NRF-NRFF15-2023-0010). SB is funded by the National Institute for Health and Care Research (NIHR) Health Protection Research Unit in Modelling and Health Economics, a partnership between UK Health Security Agency, Imperial College London and LSHTM (grant code NIHR200908). SB acknowledges: support from the Novo Nordisk Foundation via The Novo Nordisk Young Investigator Award (NNF20OC0059309); the Danish National Research Foundation (DNRF160) through the chair grant, and support from The Eric and Wendy Schmidt Fund For Strategic Innovation via the Schmidt Polymath Award (G-22-63345). DJL acknowledges funding from the Wellcome Trust for the Vaccine Impact Modelling Consortium (VIMC) Climate Change Research Programme (grant ID: 226727\_Z\_22\_Z).

## Disclaimer

“The views expressed are those of the author(s) and not necessarily those of the NIHR, UK Health Security Agency or the Department of Health and Social Care”.

## Authors contributions

Conceptualisation: ND, DJL

Data acquisition and cleaning: ND, SM, WRH, SB, DJL

Formal analysis: ND, DJL

Writing – first draft: ND, DJL

Writing – review and editing: ND, SM, WRH, SB, DJL

Supervision: DJL

## Supplementary Materials

### Supplementary Methods: Model alternatives

We considered several alternatives to the main model. Firstly, we considered a model in which vaccination effectiveness varied by dose and variant, and thus *VE*_*dv*_ is the vaccination effectiveness against SARS-CoV-2 transmission for variant *v* in a population 100% vaccinated with a dose *d*. This model, termed “Variant-specific-VE”, is given by:

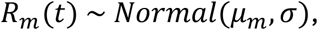

Where

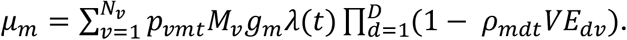

Secondly, we considered a model using the age-group-proportion of vaccinated individuals, as opposed to using the proportion of vaccinees among the overall LTLA population. In this model alternative, we defined *ρ*_*mdat*_ as the proportion of vaccinated individuals in an age group *a* = 1, …, *A* in each LTLA *m* at a timepoint *t* with a dose *d*, and *P*_*a*_ as the overall proportion of individuals in age group *a*. We only considered three age groups (15-49, 50-69 and 70-plus year olds) and thus *A* = 3. This model, termed “Age-model”, is given by:

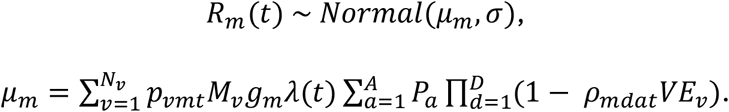

Finally, we considered a model alternative to the above in which vaccination effectiveness varied by dose, variant and age group. *VE*_*adv*_ is the vaccination effectiveness against SARS-CoV-2 transmission for variant *v* in a population 100% vaccinated with a dose *d* for age group *a*. This model, termed “Age-variant-specific-VE”, is given by:

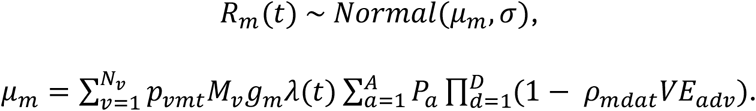

Due to data on the proportion of vaccinees over time across LTLAs by age group being less complete, these last two models reduce to our main model.

Estimates of vaccine effectiveness from each of these model variants are shown in Figure S13.

## Supplementary Figures

**Figure S1:**
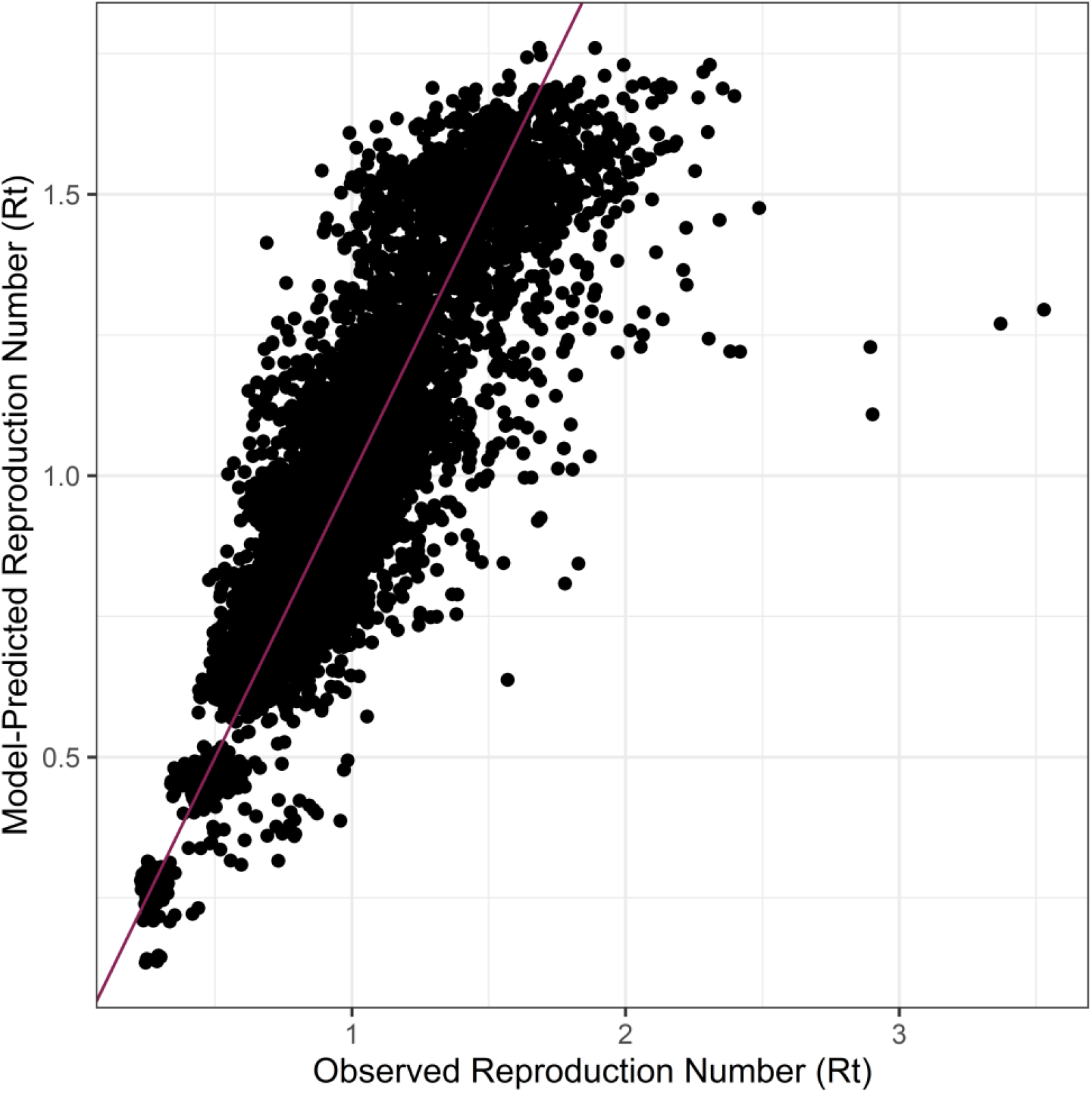
A) Observed R_t_ and model-predicted R_t_. Abbreviations: R_t_, Reproduction Number.

**Figure S2:**
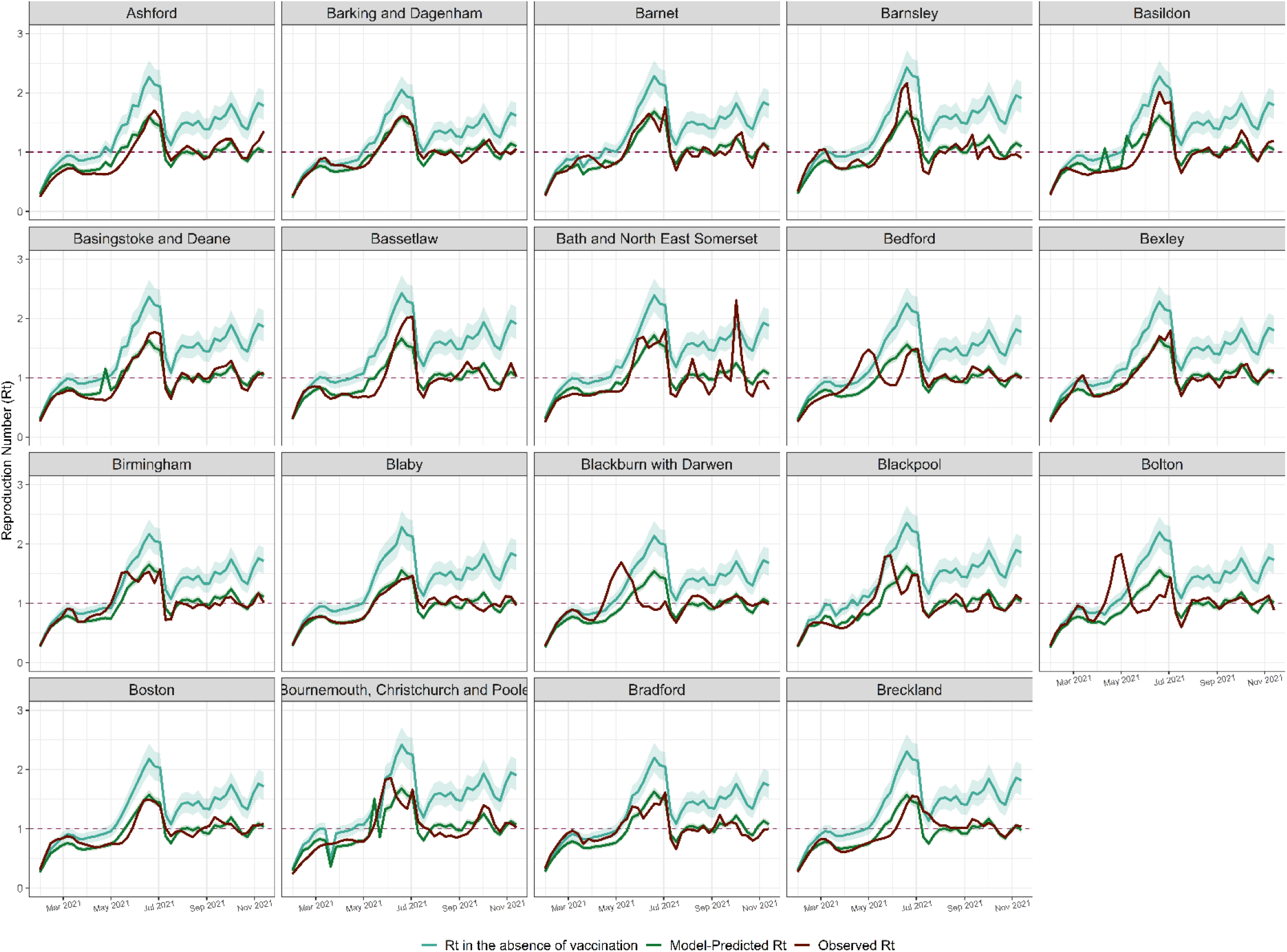
Model fits for individual LTLA: R_t_ in the absence of vaccination, model-predicted and observed R_t_ in 2021. Each LTLA R_t_ in the absence of vaccination, model-predicted R_t_ and observed R_t_ are plotted as a continuous line. The 2.5% and 97.5% percentiles for each LTLA R_t_ in the absence of vaccination and model-predicted R_t_ are plotted as a ribbon line. Abbreviations: LTLA, Lower Tier Local Authority; R_t_, Reproduction Number.

**Figure S3:**
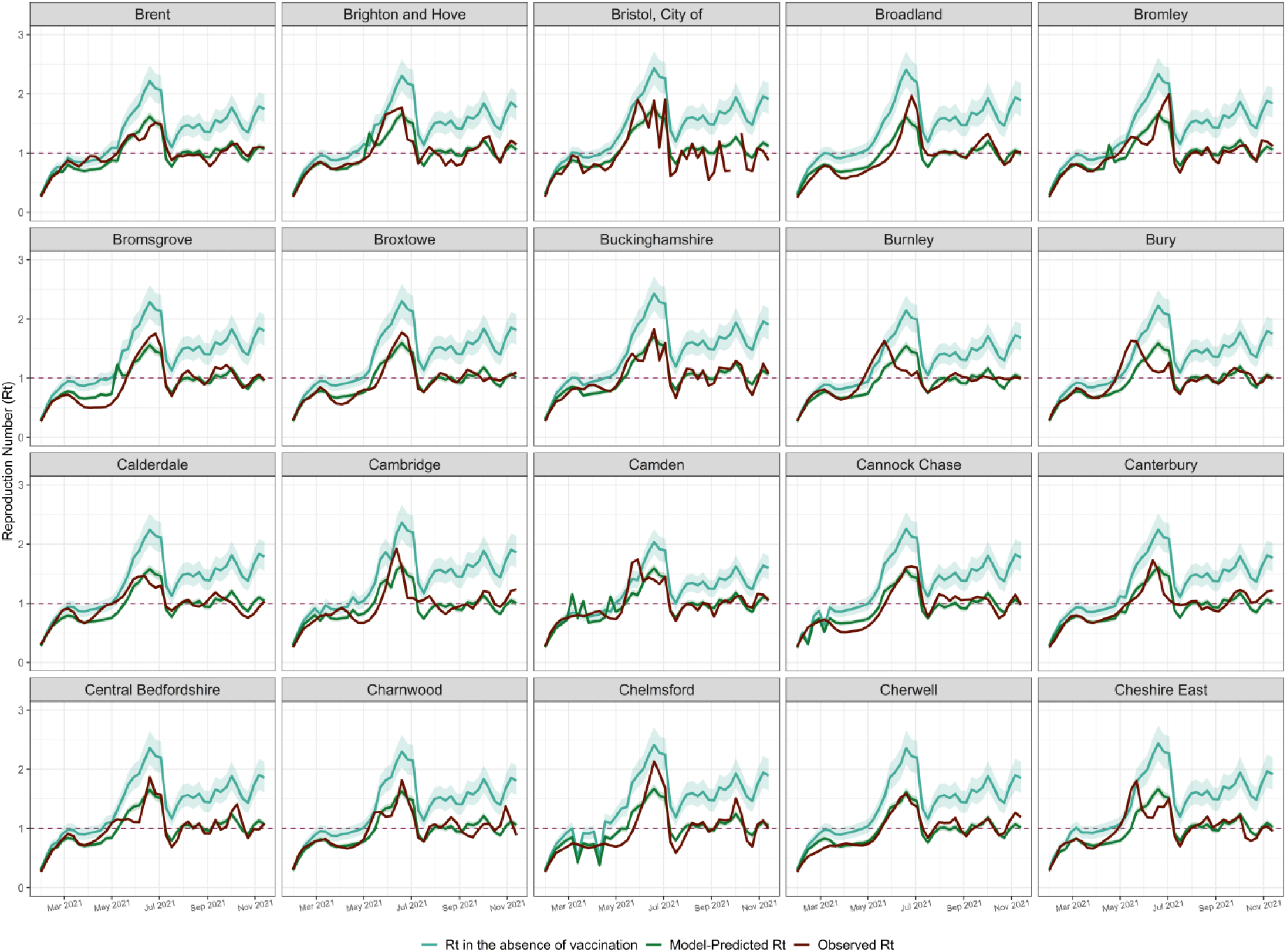
Model fits for individual LTLA: R_t_ in the absence of vaccination, model-predicted and observed R_t_ in 2021. Each LTLA R_t_ in the absence of vaccination, model-predicted R_t_ and observed R_t_ are plotted as a continuous line. The 2.5% and 97.5% percentiles for each LTLA R_t_ in the absence of vaccination and model-predicted R_t_ are plotted as a ribbon line. Abbreviations: LTLA, Lower Tier Local Authority; R_t_, Reproduction Number.

**Figure S4:**
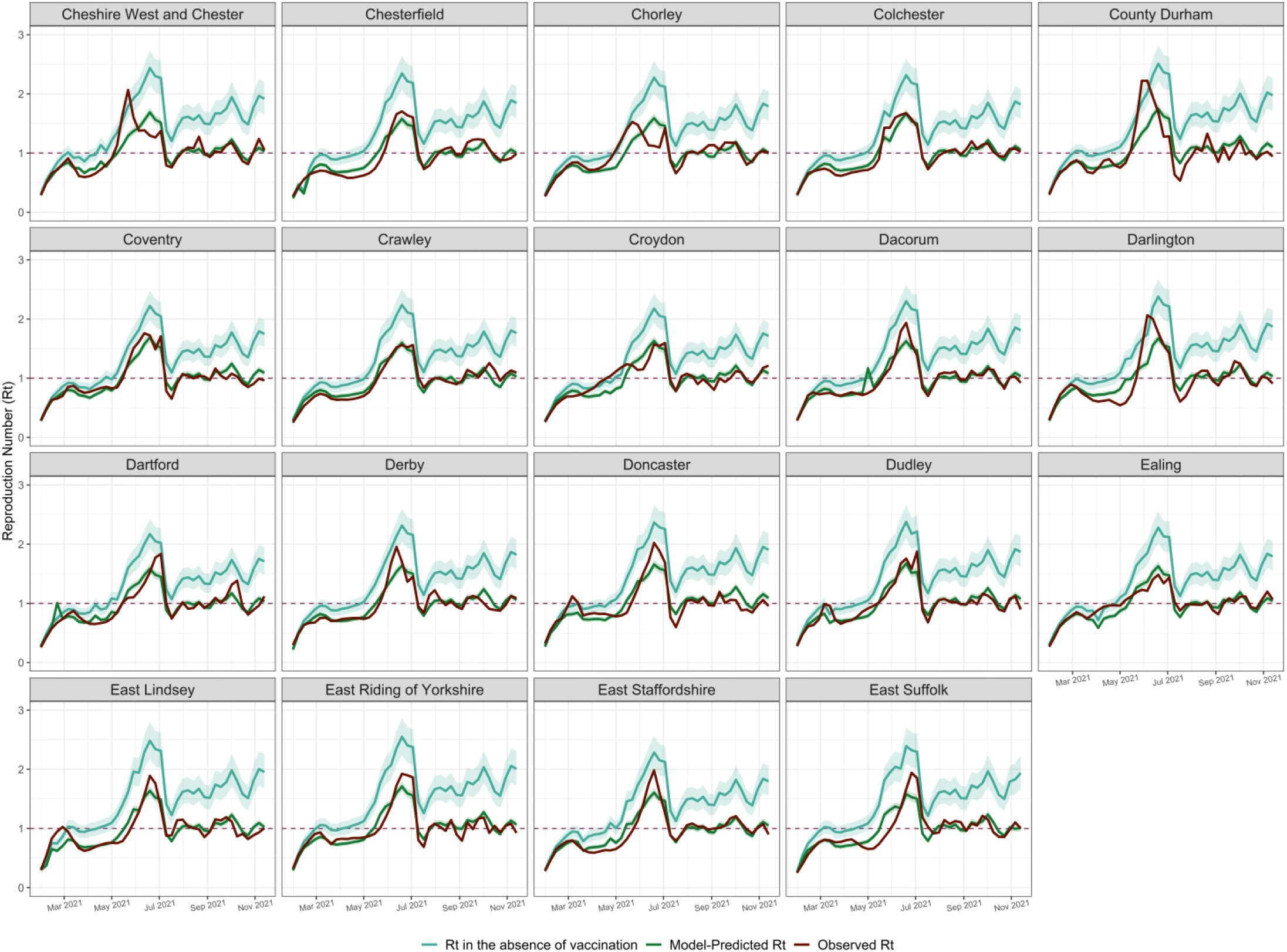
Model fits for individual LTLA: R_t_ in the absence of vaccination, model-predicted and observed R_t_ in 2021. Each LTLA R_t_ in the absence of vaccination, model-predicted R_t_ and observed R_t_ are plotted as a continuous line. The 2.5% and 97.5% percentiles for each LTLA R_t_ in the absence of vaccination and model-predicted R_t_ are plotted as a ribbon line. Abbreviations: LTLA, Lower Tier Local Authority; R_t_, Reproduction Number.

**Figure S5:**
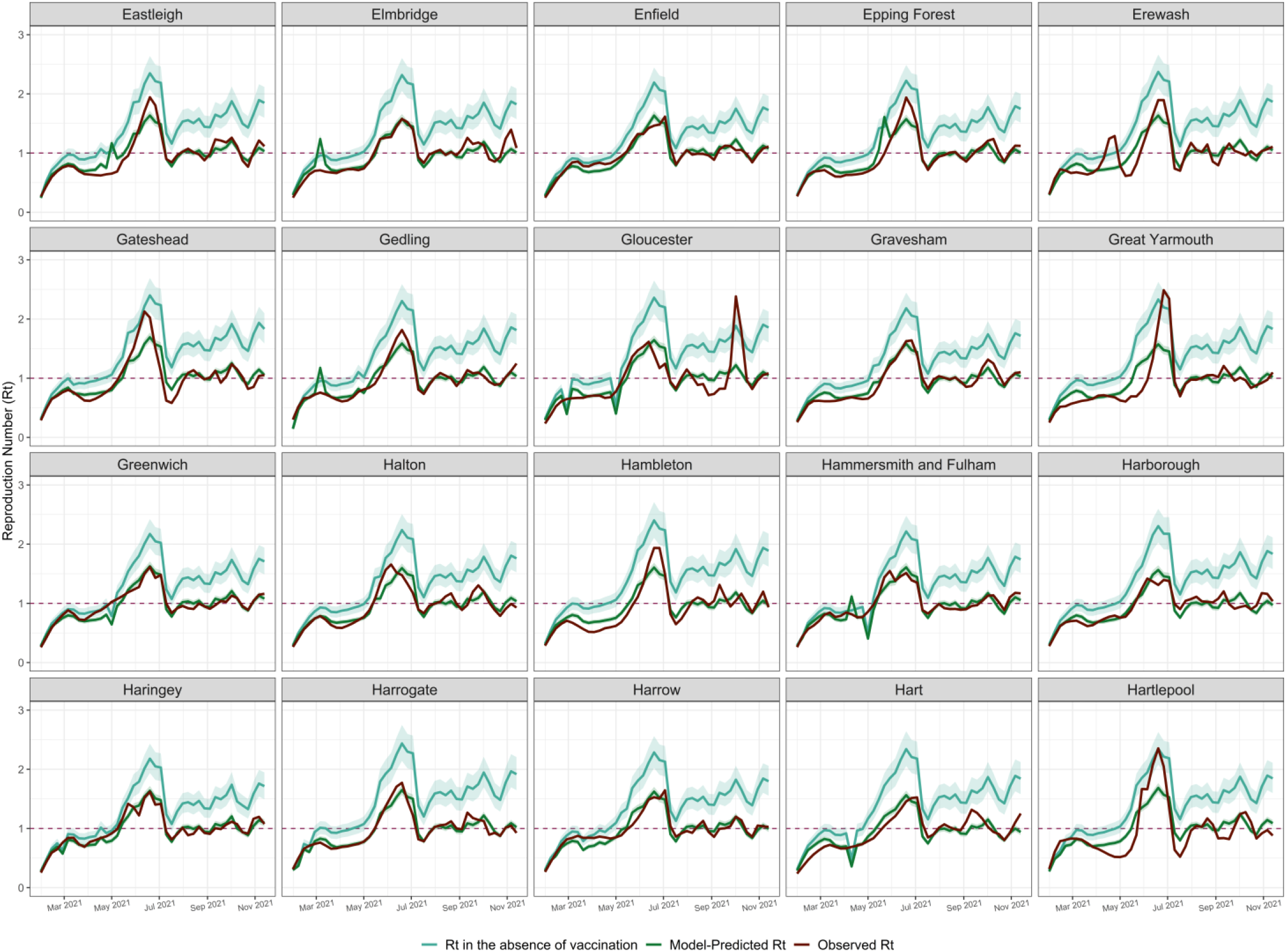
Model fits for individual LTLA: R_t_ in the absence of vaccination, model-predicted and observed R_t_ in 2021. Each LTLA R_t_ in the absence of vaccination, model-predicted R_t_ and observed R_t_ are plotted as a continuous line. The 2.5% and 97.5% percentiles for each LTLA R_t_ in the absence of vaccination and model-predicted R_t_ are plotted as a ribbon line. Abbreviations: LTLA, Lower Tier Local Authority; R_t_, Reproduction Number.

**Figure S6:**
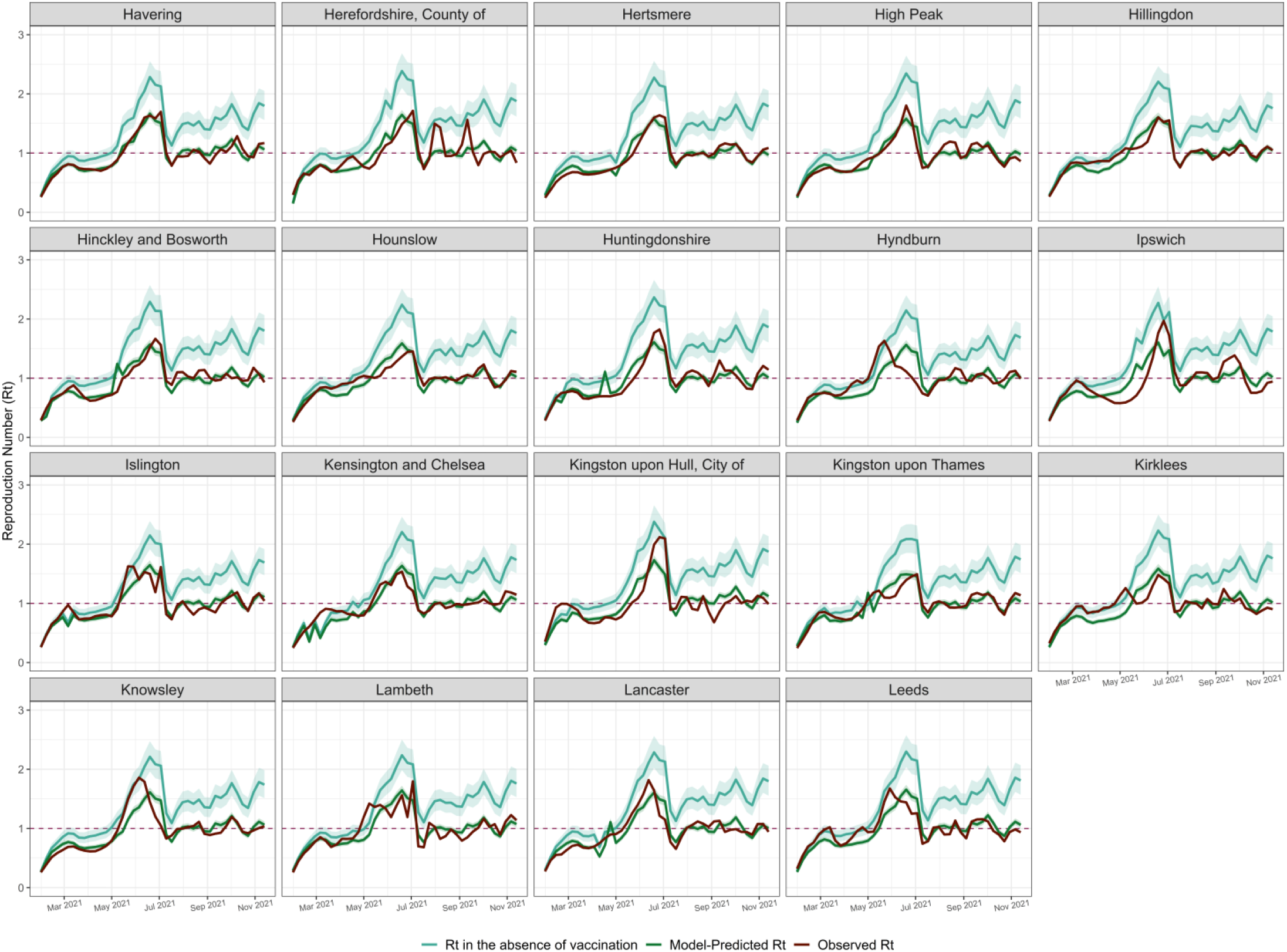
Model fits for individual LTLA: R_t_ in the absence of vaccination, model-predicted and observed R_t_ in 2021. Each LTLA R_t_ in the absence of vaccination, model-predicted R_t_ and observed R_t_ are plotted as a continuous line. The 2.5% and 97.5% percentiles for each LTLA R_t_ in the absence of vaccination and model-predicted R_t_ are plotted as a ribbon line. Abbreviations: LTLA, Lower Tier Local Authority; R_t_, Reproduction Number.

**Figure S7:**
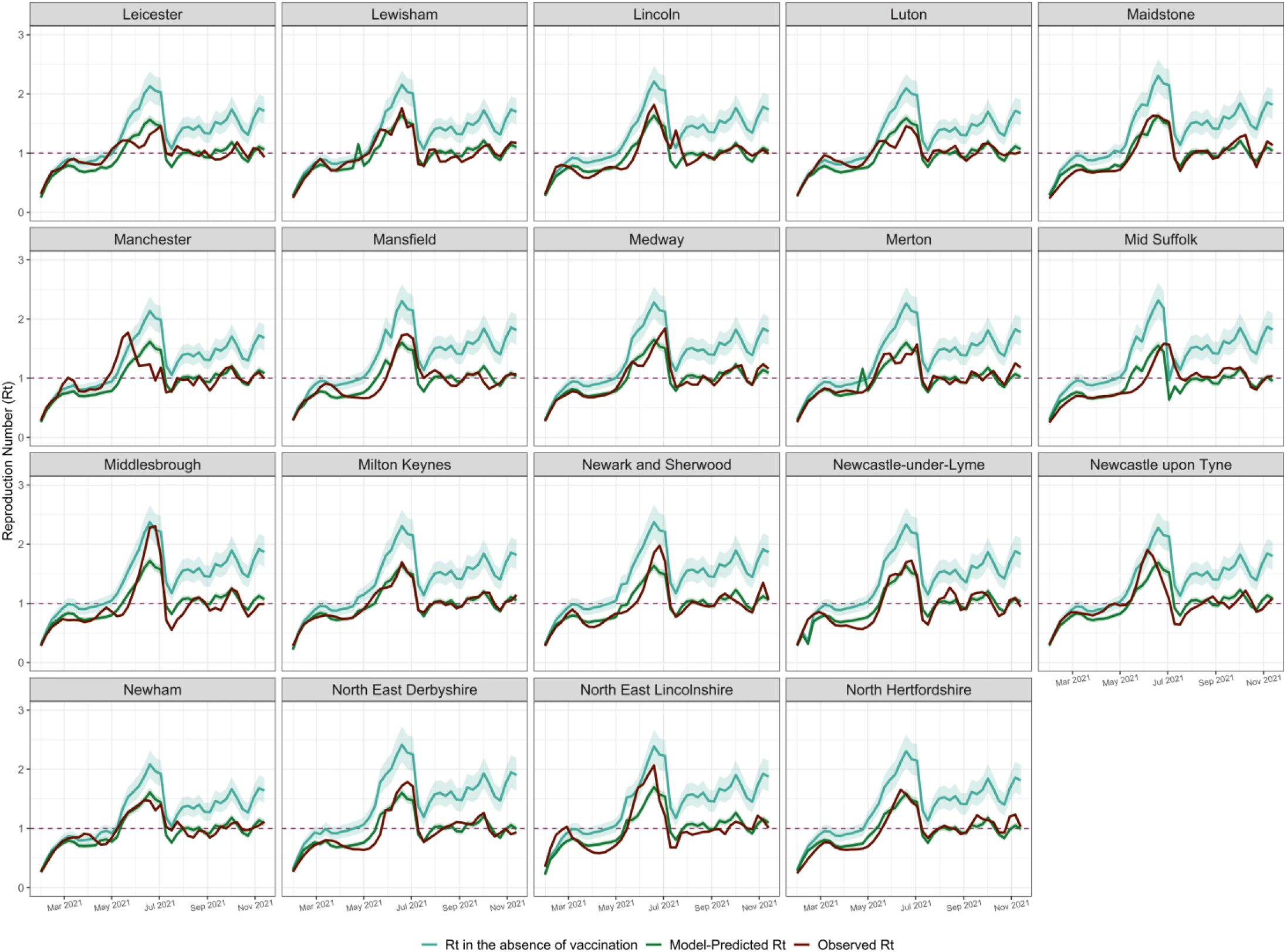
Model fits for individual LTLA: R_t_ in the absence of vaccination, model-predicted and observed R_t_ in 2021. Each LTLA R_t_ in the absence of vaccination, model-predicted R_t_ and observed R_t_ are plotted as a continuous line. The 2.5% and 97.5% percentiles for each LTLA R_t_ in the absence of vaccination and model-predicted R_t_ are plotted as a ribbon line. Abbreviations: LTLA, Lower Tier Local Authority; R_t_, Reproduction Number.

**Figure S8:**
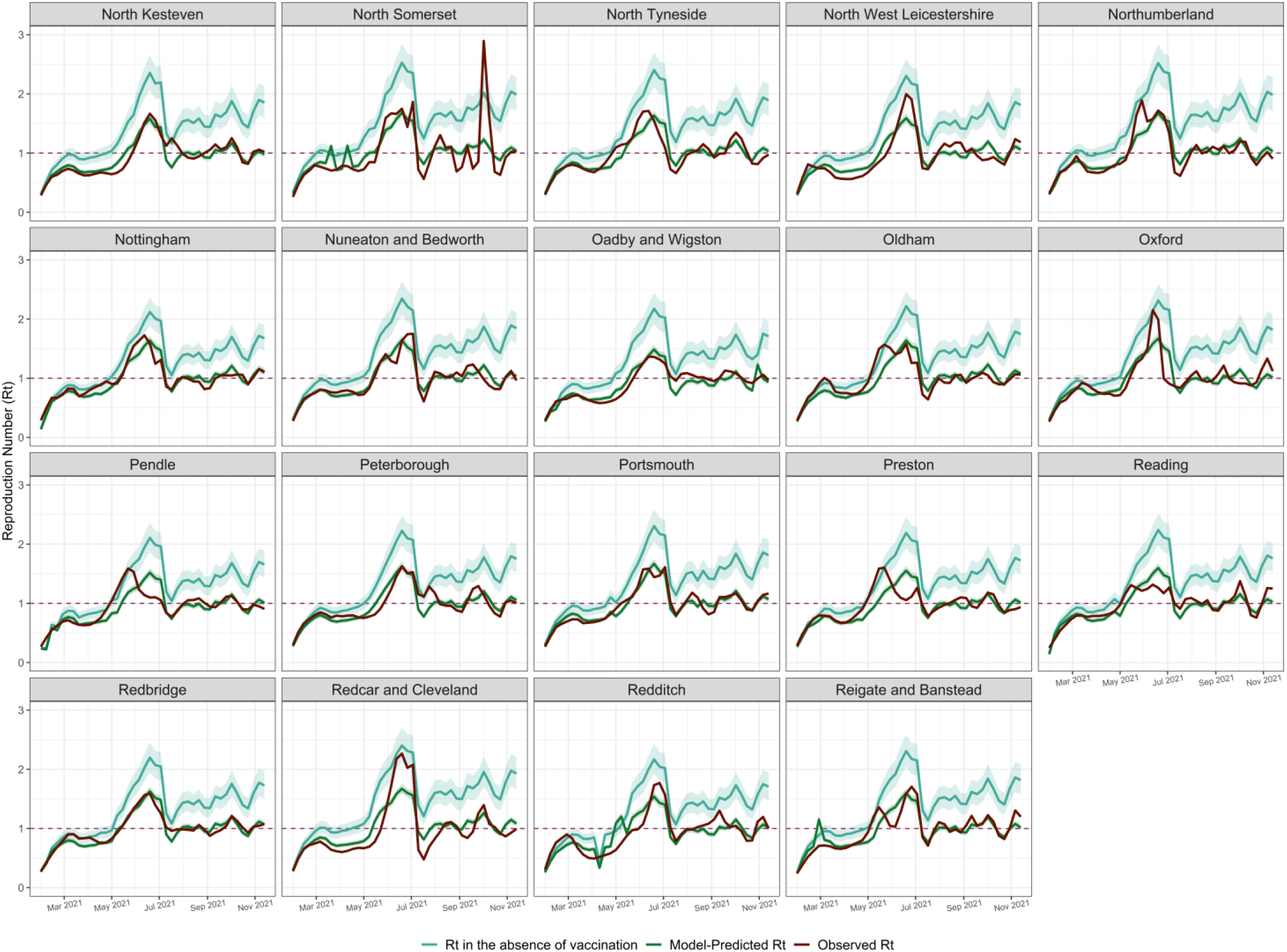
Model fits for individual LTLA: R_t_ in the absence of vaccination, model-predicted and observed R_t_ in 2021. Each LTLA R_t_ in the absence of vaccination, model-predicted R_t_ and observed R_t_ are plotted as a continuous line. The 2.5% and 97.5% percentiles for each LTLA R_t_ in the absence of vaccination and model-predicted R_t_ are plotted as a ribbon line. Abbreviations: LTLA, Lower Tier Local Authority; R_t_, Reproduction Number.

**Figure S9:**
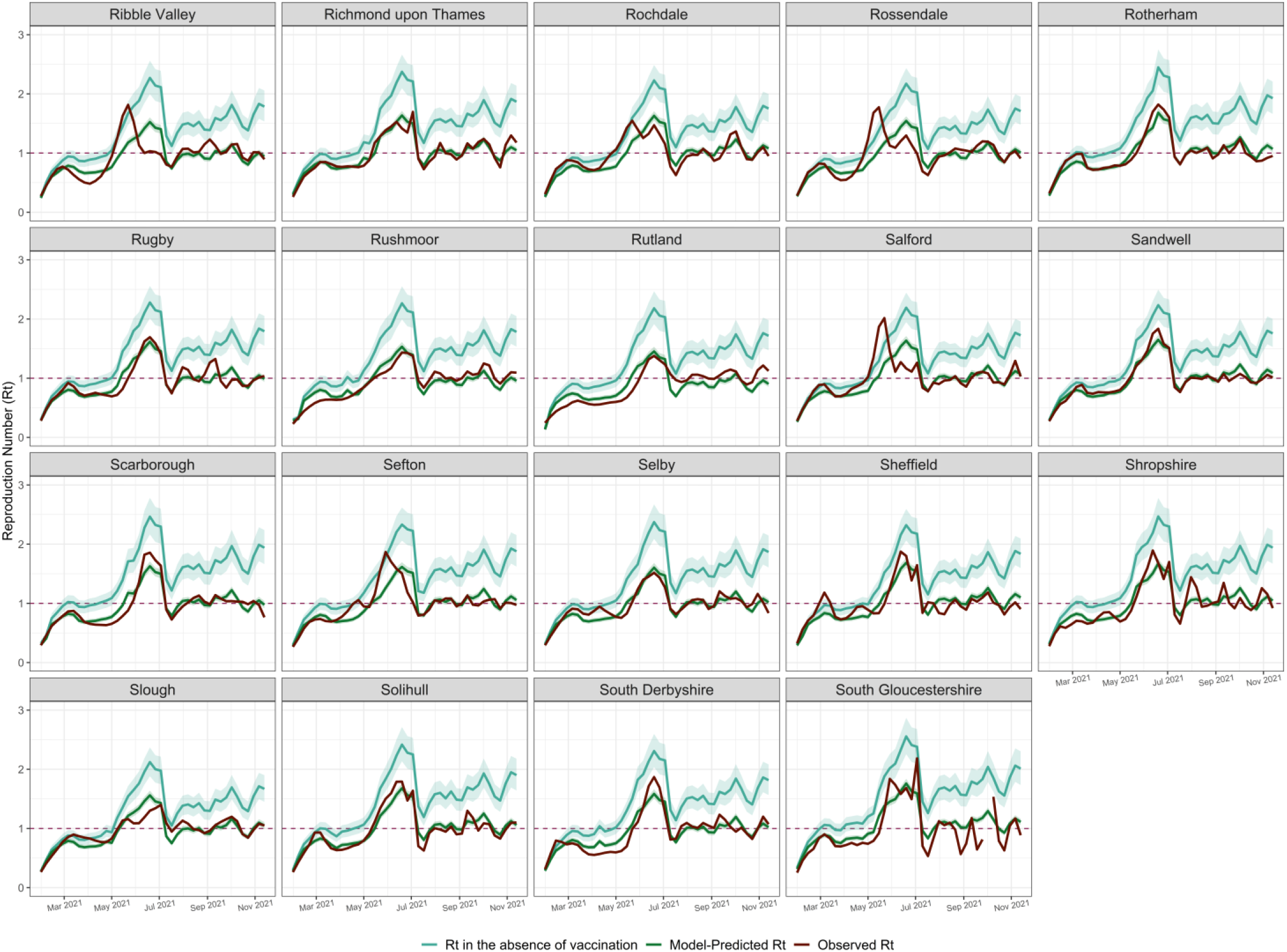
Model fits for individual LTLA: R_t_ in the absence of vaccination, model-predicted and observed R_t_ in 2021. Each LTLA R_t_ in the absence of vaccination, model-predicted R_t_ and observed R_t_ are plotted as a continuous line. The 2.5% and 97.5% percentiles for each LTLA R_t_ in the absence of vaccination and model-predicted R_t_ are plotted as a ribbon line. Abbreviations: LTLA, Lower Tier Local Authority; R_t_, Reproduction Number.

**Figure S10:**
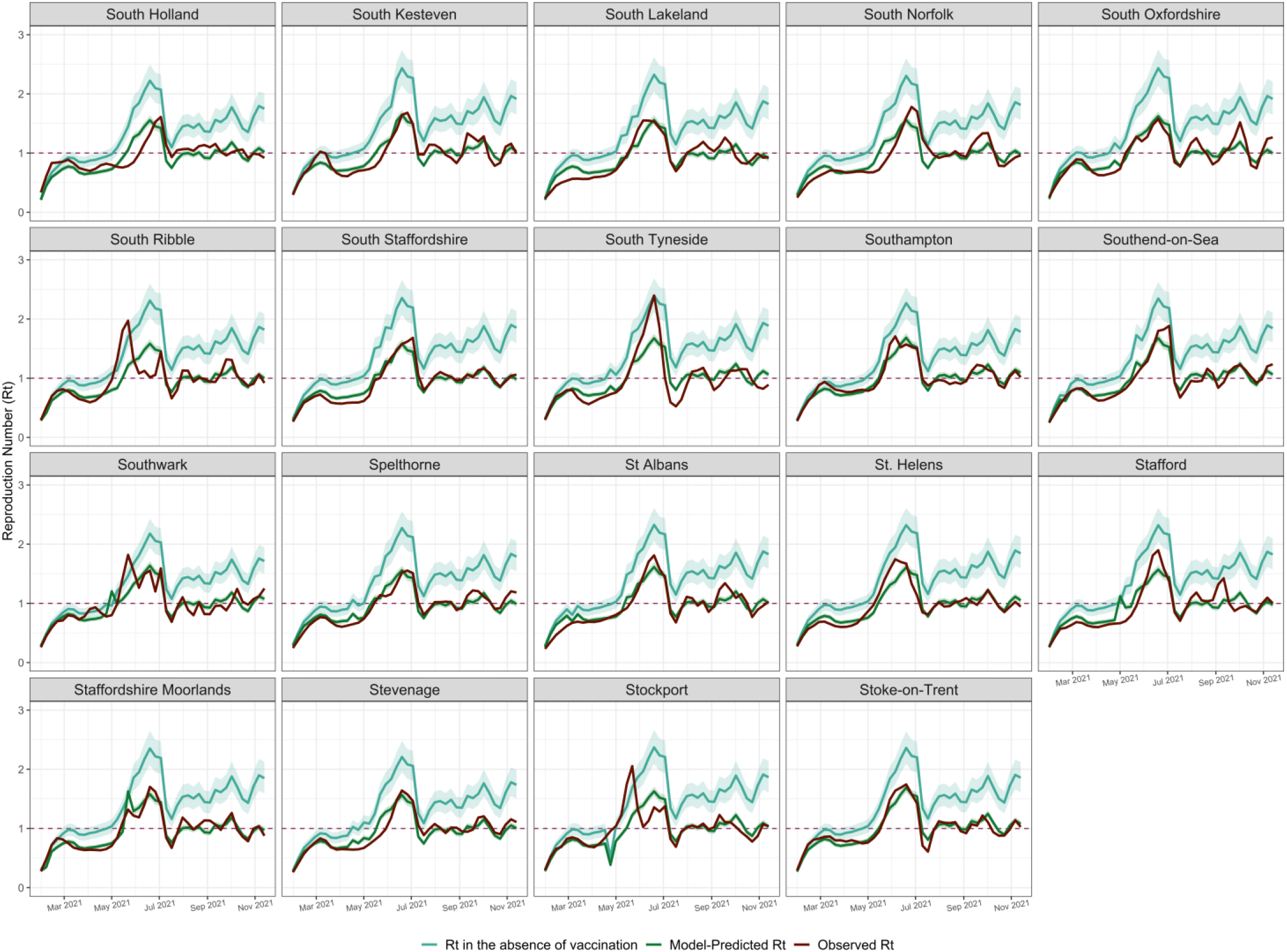
Model fits for individual LTLA: R_t_ in the absence of vaccination, model-predicted and observed R_t_ in 2021. Each LTLA R_t_ in the absence of vaccination, model-predicted R_t_ and observed R_t_ are plotted as a continuous line. The 2.5% and 97.5% percentiles for each LTLA R_t_ in the absence of vaccination and model-predicted R_t_ are plotted as a ribbon line. Abbreviations: LTLA, Lower Tier Local Authority; R_t_, Reproduction Number.

**Figure S11:**
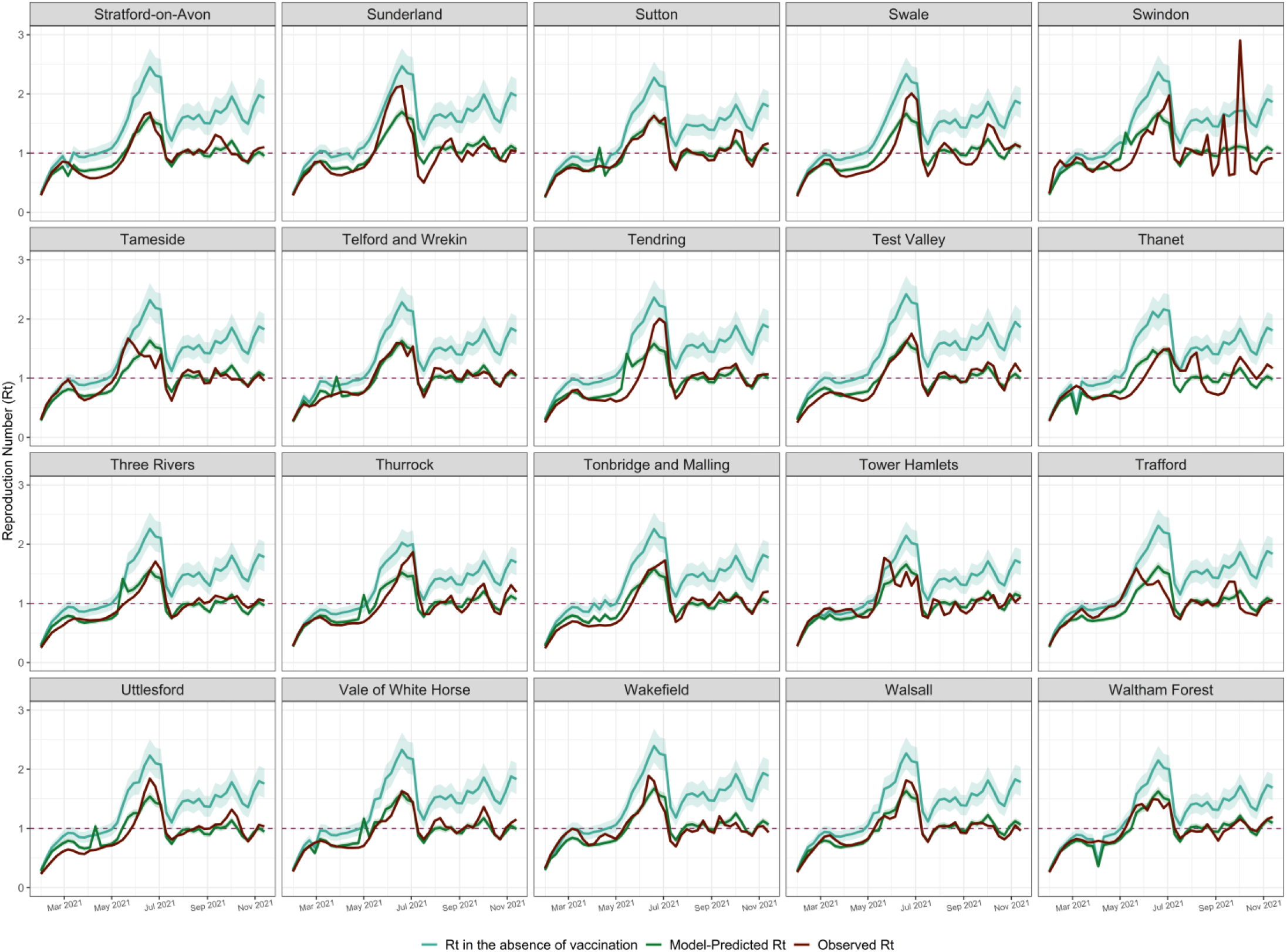
Model fits for individual LTLA: R_t_ in the absence of vaccination, model-predicted and observed R_t_ in 2021. Each LTLA R_t_ in the absence of vaccination, model-predicted R_t_ and observed R_t_ are plotted as a continuous line. The 2.5% and 97.5% percentiles for each LTLA R_t_ in the absence of vaccination and model-predicted R_t_ are plotted as a ribbon line. Abbreviations: LTLA, Lower Tier Local Authority; R_t_, Reproduction Number.

**Figure S12:**
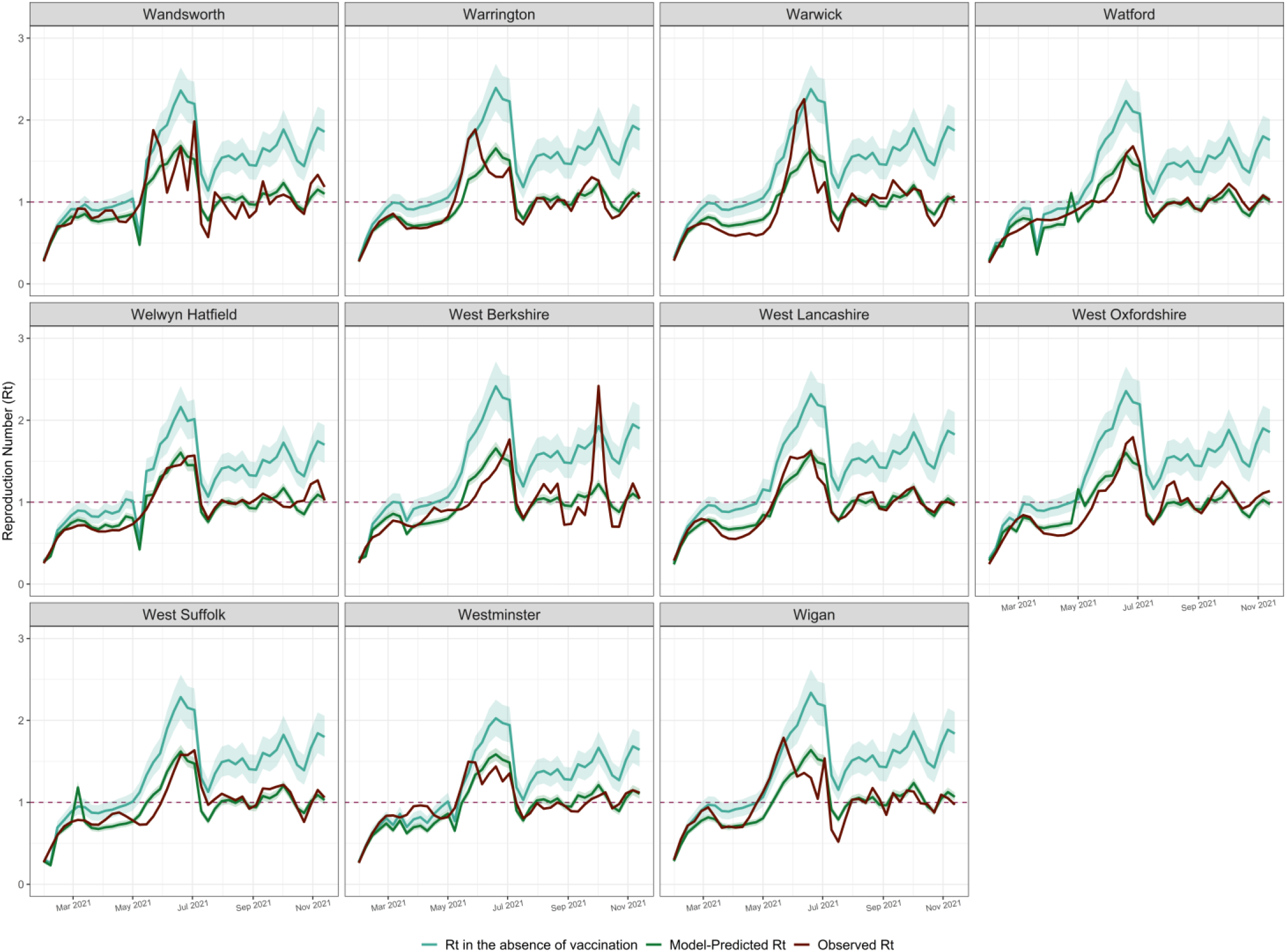
Model fits for individual LTLA: R_t_ in the absence of vaccination, model-predicted and observed R_t_ in 2021. Each LTLA R_t_ in the absence of vaccination, model-predicted R_t_ and observed R_t_ are plotted as a continuous line. The 2.5% and 97.5% percentiles for each LTLA R_t_ in the absence of vaccination and model-predicted R_t_ are plotted as a ribbon line. Abbreviations: LTLA, Lower Tier Local Authority; R_t_, Reproduction Number.

**Figure S13:**
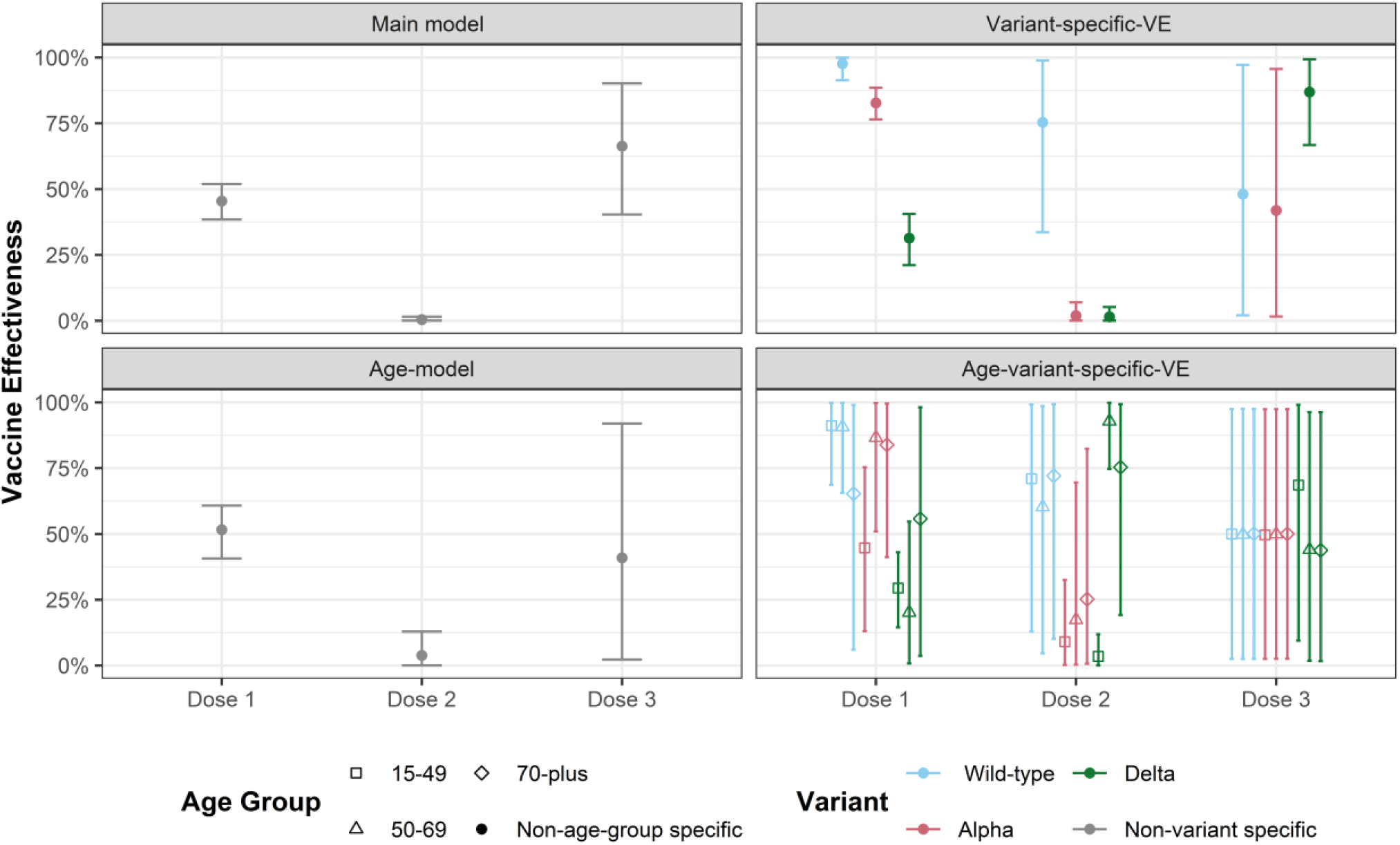
Vaccine effectiveness across model variants. Model predictions for vaccine effectiveness (A) are shown as point estimates depicting the average across iterations, with error bars showing 2.5% percentile 97.5% percentile.

## Supplementary Tables

**Table S1:**
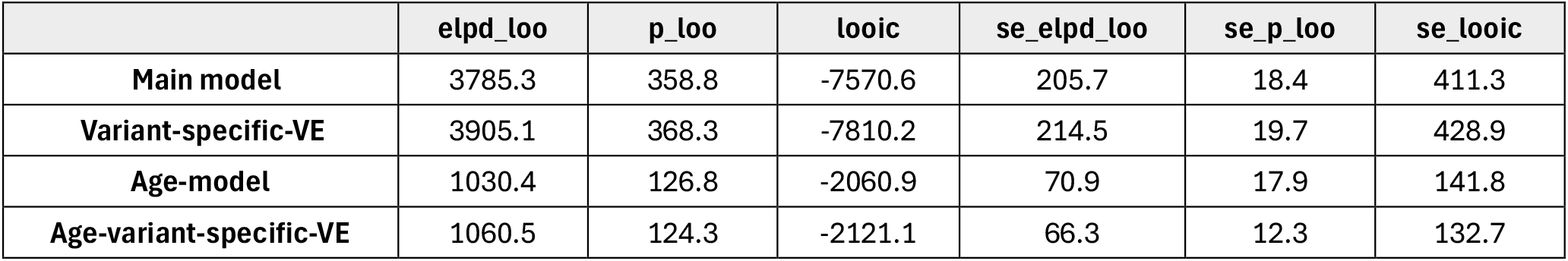
Leave-one-out cross validation across model variants.

